# Alexithymia moderates brain function during suppression of negative emotions in fibromyalgia

**DOI:** 10.1101/2025.09.12.25335623

**Authors:** Amaia Aguirre de Cárcer Vidal, Nell Norman-Nott, Sylvia M. Gustin, Yann Quidé

## Abstract

Fibromyalgia is associated with altered brain function and emotion regulation difficulties. Alexithymia is common in fibromyalgia and may further affect emotion regulation in this population. This study examined the interplay of alexithymia and fibromyalgia on the brain function during suppression of negative emotions. Twenty-seven females with fibromyalgia and 30 controls performed an emotion regulation task while undergoing functional magnetic resonance imaging. Alexithymia was assessed using the Toronto Alexithymia Scale (TAS). A series of whole-brain multiple linear regressions was performed to estimate the effects of group, alexithymia and their interaction on activation and seed-based task-related functional connectivity. Reduced activation in the left middle frontal gyrus during negative emotion suppression, as well as reduced left amygdala–right temporo-parietal junction connectivity, increased left anterior insula-right posterior insula connectivity, and increased left anterior cingulate cortex-precuneus and inferior parietal lobule connectivity were evident in fibromyalgia. Increasing alexithymia was associated to stronger left anterior insula–inferior parietal lobule connectivity, across all groups. Following significant association between the group-by-TAS interaction and right amygdala–lingual gyri connectivity, moderation analysis indicated that increasing alexithymia was associated with reduced connectivity in healthy controls but not in fibromyalgia. This study highlights disrupted integration between emotional, interoceptive, and self-referential networks as a key feature of negative emotion suppression in fibromyalgia. Alexithymia moderates these disruptions by reducing emotional clarity and increasing internal focus. Interventions enhancing interoceptive awareness, fostering emotional understanding, and strengthening cognitive regulation may help normalise neural function and improve emotional functioning, especially in individuals with elevated alexithymia.

## Introduction

Fibromyalgia is a chronic, idiopathic condition characterised by widespread musculoskeletal pain without identifiable tissue damage or inflammation (World Health Organization, 2019). Prevalence ranges from 0.2% to 6.6% (Marques et al., 2017), depending on diagnostic criteria (Fors et al., 2024). It disproportionately affects women and is frequently accompanied by depression, anxiety and emotion regulation problems (Telli & Akkus, 2025). It also impacts brain function, particularly during emotion processing and regulation (Balducci et al., 2024). Emotion regulation refers to processes used to monitor, modify, and adjust emotions in response to internal or external demands (Gross, 1998). Alexithymia, characterised by difficulties identifying, describing and processing emotions, contributes to emotion regulation problems and is commonly elevated in people with fibromyalgia, even when controlling for comorbid mood disorders (Habibi Asgarabad et al., 2023; Renzi et al., 2025; Tesio et al., 2018). To the best of our knowledge, no previous study has determined the contribution of alexithymia to the brain alterations seen in fibromyalgia during emotion regulation.

According to the Fibromyalgia Integrative Training for Self-regulation and Salience (FITSS) model, dysfunction within the salience network contributes to maladaptive emotional and somatic processing and impairments in self-regulation (Pinto et al., 2023), potentially leading to ineffective emotion regulation. The salience network, comprising the ACC, anterior insula, and amygdala, critically detect and integrate salient internal and external stimuli, coordinate the allocation of attentional and cognitive resources, and initiate behavioural responses (Uddin, 2015). The salience network is closely connected to the central executive network, made of the dorsolateral prefrontal cortex (dlPFC) and posterior parietal cortices, and involved in task-oriented, higher cognitive processes (Corbetta & Shulman, 2002; Marek & Dosenbach, 2018). Heightened connectivity between the salience and the default-mode network, made of the posterior cingulate cortex (PCC)/precuneus, medial prefrontal cortex (mPFC), bilateral temporo-parietal junctions (TPJ) and the hippocampus (Raichle, 2015), is consistently reported in fibromyalgia (Napadow et al., 2010; Pinto et al., 2023). In fibromyalgia, altered interactions among these networks are associated with problems in emotion regulation and decision-making, aberrant self-referential and interoceptive processing (Balducci et al., 2024; Johansson et al., 2024; Pinto et al., 2023).

Among the different processes involved in emotion regulation, suppression is a regulation strategy involving inhibiting outward expressions, that typically recruits prefrontal control regions to modulate limbic activity (Gross, 2002). Suppression is associated with increased cognitive demand, and its neural effects differ in individuals with heightened interoceptive focus or dysregulation, such as those with fibromyalgia (Balducci et al., 2024; Rost et al., 2021). As fibromyalgia is associated with higher levels of alexithymia (Marchi et al., 2019), it is not surprising that similar patterns of activation and connectivity are observed in relation to alexithymia (Butera et al., 2023; Ho et al., 2016; van der Velde et al., 2015). Thus, alexithymia may influence brain function associated with emotion regulation, especially suppression of negative emotions. However, no study to date has investigated this relationship in fibromyalgia.

The current study examined brain activation and functional connectivity during the suppression of negative emotions in women with fibromyalgia compared to healthy controls, and how alexithymia moderates these neural responses. Compared to controls, women with fibromyalgia were expected to show increased activation and connectivity among regions core to the salience network (ACC, anterior insula, amygdala), and stronger coupling with executive control (e.g., dlPFC) and default mode networks (e.g., mPFC) involved in emotion processing and regulation. Higher alexithymia scores were expected to be associated with greater limbic hyper-reactivity and reduced prefrontal regulation within these networks.

## Material and Methods

### Participants

Data from 33 females with fibromyalgia and 33 females without a chronic pain condition (Controls) were accessed from the publicly available OpenNeuro dataset ds004144 (https://openneuro.org/datasets/ds004144/versions/1.0.2) (Balducci et al., 2022b). Recruitment and clinical assessment details are available elsewhere (Balducci et al., 2022a). Participants were adults (18-50 years old) with at least elementary education. Diagnoses of fibromyalgia were given by a rheumatologist or an internal medicine specialist, confirmed by the American College of Rheumatology 1990 (Wolfe et al., 1990) and 2016 criteria (Wolfe et al., 2016). Exclusion criteria included the presence of a major psychiatric, cardiovascular, neurological or other pain conditions (see details in (Balducci et al., 2022a)). All participants answered a standardized instrument to measure socio-economic level in Mexican households, the Socio-economic levels Questionnaire of the Mexican Association of Market Research and Public Opinion Agencies Questionnaire (AMAI NSE 8×7). Details are presented Table 1. The protocol was approved by the Research Ethics Committee of the National Institute of Psychiatry “Ramón de la Fuente Muñiz” in Mexico City, in accordance with the Declaration of Helsinki, and all participants provided written informed consent.

**Table 1.**
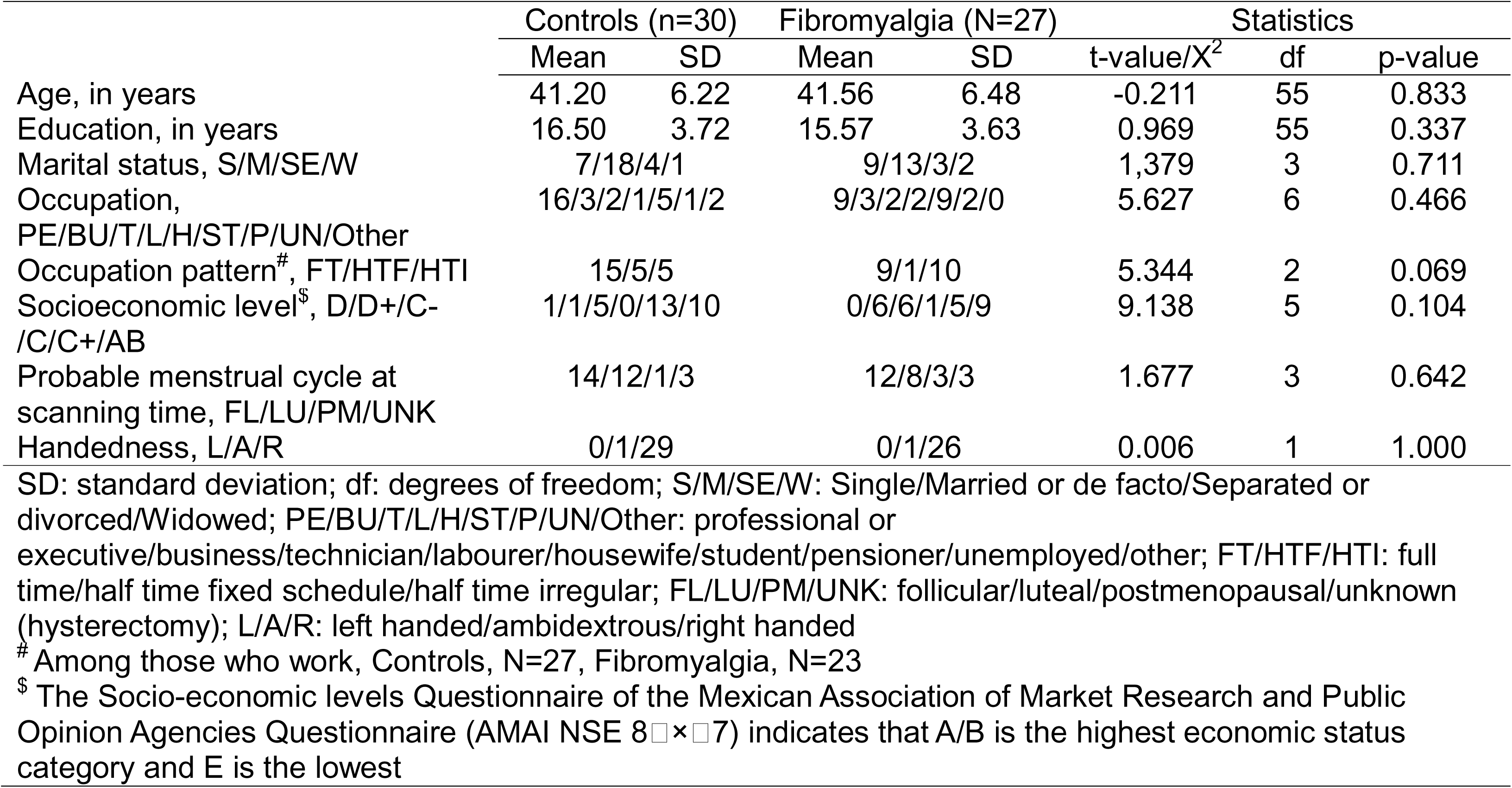
Socio-demographic characteristics of the sample

|  | Controls (n=30) |  | Fibromyalgia (N=27) |  | Statistics |  |  |
| --- | --- | --- | --- | --- | --- | --- | --- |
|  | Mean | SD | Mean | SD | t-value/X<sup>2</sup> | df | p-value |
| Age, in years | 41.20 | 6.22 | 41.56 | 6.48 | -0.211 | 55 | 0.833 |
| Education, in years | 16.50 | 3.72 | 15.57 | 3.63 | 0.969 | 55 | 0.337 |
| Marital status, S/M/SE/W | 7/18/4/1 |  | 9/13/3/2 |  | 1,379 | 3 | 0.711 |
| Occupation, PE/BU/T/L/H/ST/P/UN/Other | 16/3/2/1/5/1/2 |  | 9/3/2/2/9/2/0 |  | 5.627 | 6 | 0.466 |
| Occupation pattern<sup>#</sup>, FT/HTF/HTI | 15/5/5 |  | 9/1/10 |  | 5.344 | 2 | 0.069 |
| Socioeconomic level<sup>\$</sup>, D/D+/C-/C/C+/AB | 1/1/5/0/13/10 | | 0/6/6/1/5/9 | | 9.138 | 5 | 0.104 |
| Probable menstrual cycle at scanning time, FL/LU/PM/UNK | 14/12/1/3 |  | 12/8/3/3 |  | 1.677 | 3 | 0.642 |
| Handedness, L/A/R | 0/1/29 |  | 0/1/26 |  | 0.006 | 1 | 1.000 |

SD: standard deviation; df: degrees of freedom; S/M/SE/W: Single/Married or de facto/Separated or divorced/Widowed; PE/BU/T/L/H/ST/P/UN/Other: professional or executive/business/technician/labourer/housewife/student/pensioner/unemployed/other; FT/HTF/HTI: full time/half time fixed schedule/half time irregular; FL/LU/PM/UNK: follicular/luteal/postmenopausal/unknown (hysterectomy); L/A/R: left handed/ambidextrous/right handed

### Clinical scales

No longer than two weeks prior to the imaging session, participants completed a comprehensive clinical and psychological assessment. The current health status and symptom severity associated with fibromyalgia were assessed using the 20-item Spanish version of the Fibromyalgia Impact Questionnaire (FIQ) (Vargas-Alarcon et al., 2007). Pain from a dimensional perspective was evaluated using the 78-item Spanish version of the McGill pain questionnaire (Masedo & Esteve, 2000). Duration and current treatment for fibromyalgia were recorded using the Fibromyalgia General Questionnaire.

### Other clinical scales

Levels of alexithymia were measured using the 20 items self-rated Toronto Alexithymia Scale (TAS) (Bagby et al., 1994). Although subdomains of alexithymia can be derived from the TAS (difficulty identifying emotions, difficulty expressing emotions and externally oriented thought), only the total TAS score was used for focal analyses.

Severity of depressive symptoms were evaluated using the Spanish version of the Hamilton Depression Rating Scale (HAMD) (Carrozzino et al., 2020). The version used comprised 21 items, each item being scored on a Likert-type scale (scores ranging 0–4). The Spanish version of the Hamilton Anxiety Rating Scale (HAMA) (Bruss et al., 1994) was used to measure the severity of anxiety symptoms experienced. This 14-item Likert-type scale scored the intensity, frequency and dysfunction caused by symptoms (scores ranging 0–4).

### Functional magnetic resonance imaging task

Participants performed a validated emotion processing and regulation task (Balducci et al., 2024; van Kleef et al., 2022). The task required the participants to respond to three different instructions (attend, reappraise, suppress) and for three emotionally valenced stimuli (neutral, positive, negative pictures from the International Affective Picture System) (Lang et al., 2005) for a total of seven conditions (attend neutral, attend negative, attend positive, reappraise negative, reappraise positive, suppress negative, suppress positive). The task was implemented in a block design, with each block representing each condition, and each condition was repeated three times (three blocks/condition). Each block contained the instruction for that block (attend, increase, decrease or suppress), followed by four pictures, and three evaluation screens with a visual analogue scale on each. Blocks were separated by a fixation cross of varying length (8000 to 14500 ms). Using the visual analogue scales (ranging 0–10) participants rated the intensity and valence of their current arousal (How awake do you feel at this moment?”), affective state (“How do you feel at this moment?”), and the physical pain at that moment (“How intense is the pain at this moment?”). Blocks were presented pseudorandomized (first a neutral block, then six emotional blocks in random order, then seven blocks including both emotional and neutral blocks in random order, then six emotional blocks in random order, and finally a neutral block). This study focused solely on the suppression of negative emotion blocks.

### Magnetic resonance imaging

#### Images acquisition and processing

Whole-brain T1-weighted and T2*-weighted scans were acquired using a Philips 3T Ingenia scanner (Philips Healthcare, Best, The Netherlands) with a 32-channel phased array coil. As described elsewhere (Balducci et al., 2024; Balducci et al., 2022a), scanning parameters for the three-dimensional T1-weighted FFE SENSE sequence were: repetition time (TR) = 7.0 ms, echo time (TE) = 3.5 ms, flip angleL=L8°, field of view (FOV)L=L240Lmm2, matrixL=L240L×L240Lmm, number of slicesL=L180, gapL=L0, planeL=Lsagittal, slice thicknessL=L1.0 mm, voxel sizeL=L1L×L1L×L1Lmm. Scanning parameters for the emotion regulation task sequence were: gradient-echo EPI, TR/TEL=L2000/30.001Lms, FAL=L75°, matrixL=L80L×L80, FOVL=L240 mm^2^, voxel sizeL=L3L×L3L×L3Lmm, slice thicknessL=L3.0Lmm, slice acquisition orderL=Linterleaved (ascending), number of slicesL=L36, phase encoding directionL=LAP, volumesL=L834, total task durationL=L27.8Lminutes. Image processing followed the CONN toolbox’s default pipeline (release 22.v2407; https://web.conn-toolbox.org) (Whitfield-Gabrieli and Nieto-Castanon 2012) for SPM12 (v7771; Wellcome Trust Centre for Neuroimaging, London, UK; http://www.fil.ion.ucl.ac.uk/spm) in Matlab r2024a (Mathworks Inc., Sherborn, MA, USA). See details in Supplementary Material. Following preprocessing, five participants with fibromyalgia and three controls were excluded because of excessive movements (over 10% of the total number of acquisitions identified as outliers, that is with signal intensity z-threshold > 9 and/or movements > 0.9 mm) and stimuli onset times were missing for one fibromyalgia participant. The final sample consisted of 27 females with fibromyalgia and 30 controls.

#### Seed regions

Partly consistent with (Balducci et al., 2024) we selected the left and right amygdalae, anterior insulae and ACC, core nodes of the salience network, as seed regions for this study. The amygdala seed regions (left: 220 voxels; right: 248 voxels) were extracted from the *Automated Anatomical Labeling* atlas (*aal)* (Tzourio-Mazoyer et al., 2002) available with the Marsbar toolbox (v0.45) (Brett et al., 2002). The anterior insula seed regions were extracted from the Hammers atlas (Faillenot et al., 2017; Hammers et al., 2003), with the left anterior insula seed made of regions number 86 and 92 (792 voxels) and the right anterior insula seed made of the regions number 87 and 93 (809 voxels). Finally, as in (Balducci et al., 2024), 5mm spheres around published coordinates (MNI [±6,36,10]; 81 voxels) were created for the ACC seed regions.

#### Task related functional connectivity

The generalized Psycho-Physiological Interactions toolbox (gPPI, v7.12, https://www.nitrc.org/projects/gppi) (McLaren et al., 2012) was used to estimate whole-brain task-related functional connectivity during the suppression of negative emotions for the six seed regions.

### Statistical analyses

Due to the relatively small sample size, group analyses were performed using the Statistical non-Parametric Mapping toolbox (SnPM13.1.09; http://www.nisox.org/Software/SnPM13/) for SPM12 (Nichols & Holmes, 2002). This toolbox uses permutation tests, avoiding normality assumptions and reducing false positives in small samples (Eklund et al., 2016; Woo et al., 2014). Multiple linear regressions tested the main effects of group (controls versus fibromyalgia), levels of alexithymia (TAS total) and their interaction (the product of group × mean-centred TAS total score) on brain activation and functional connectivity during suppression of negative emotions (one model per seed). SnPM13 default parameters were used to define the cluster-forming threshold, and family-wise error correction was applied to the cluster statistics (*pFWEc* < 0.05). Cluster-based parameters were set to perform 10,000 permutations, and variance smoothing was not applied (set to [0,0,0]).

Following whole-brain analyses, raw signal at the peak of significant clusters were extracted for further analyses using R (version 4.4.0) (R Core Team, 2024) in RStudio (version 2024.04.1+748) (Posit Team, 2024). Because of the relatively small sample size and the number of seed regions investigated, gPPI analyses were exploratory and no additional correction to account for the number of seed regions studied were applied.

In case of significant associations between the group-by-TAS interaction and brain activation or functional connectivity, moderation analyses were performed using the ‘interactions’ R package (v1.1.5) (Long, 2021) on the extracted signal at the cluster peaks. Two sets of moderation analyses were performed. In the first, the effects of TAS total score (independent variable) on brain function or connectivity (dependent variable) were tested for each group (moderator). In the second, the effects of group (independent variable) were tested at three levels of TAS total scores (moderator): at 1 standard deviation (SD) below the average TAS total score (low TAS total score), at average TAS total score, and at 1 SD above the average TAS total score (high TAS total score) (Cohen et al., 2003). The Davidson– MacKinnon correction (HC3) was used to account for heteroskedasticity (Hayes & Cai, 2007) using the R package ‘sandwich’ (v3.2.2) (Zeileis, 2004; Zeileis et al., 2020). Statistical significance was set at a threshold of *p* < 0.05. A power analysis using G*Power 3.1.9.2 (Faul et al., 2009) indicated that a minimum of 54 participants was necessary for these moderation analyses (F3,50 = 2.79, λ = 18.90), confirming that our final sample (n = 57) was of sufficient size to achieve 95% power for detecting a large effect (f^2^ = 0.35) for three predictors (group, TAS total score, interaction) at α = 0.05.

## Results

### Sample characteristics

Sociodemographic and clinical details are presented Table 1 and Table 2, respectively. The fibromyalgia and the control groups were statistically matched for age, handedness, years of education, occupation levels, socio-economical levels and probable menstrual cycles (Table 1). Compared to the control group, the fibromyalgia group showed higher levels of alexithymia, depression and anxiety symptoms (Table 2).

**Table 2.** Clinical characteristics of the sample

|  | Controls (n=30) |  | Fibromyalgia (N=27) |  | Statistics |  |  |
| --- | --- | --- | --- | --- | --- | --- | --- |
|  | Mean | SD | Mean | SD | t-value/X<sup>2</sup> | df | p-value |
| Time since diagnosis | - | - | 4.10 | 4.75 | - | - | - |
| Time since symptoms | - | - | 8.26 | 10.69 | - | - | - |
| Current medication received – Yes/No | - | - | 24/3 |  | - | - | - |
| Current medication received – Number of medications used daily 0/1/2/3/4/5 | - | - | 8/13/5/1/0/0 |  | - | - | - |
| Pain during interview (VAS)<sup>\$</sup> | 1.30 | 3.56 | 44.26 | 20.09 | **-10.957** | **27.464** | **<0.001** |
| FIQ total score | - | - | 32.01 | 9.58 | - | - | - |
| McGill Pain questionnaire – sensory dimension | - | - | 42.92 | 12.53 | - | - | - |
| McGill Pain questionnaire – affective dimension | - | - | 7.38 | 3.55 | - | - | - |
| McGill Pain questionnaire – evaluative dimension | - | - | 3.23 | 1.77 | - | - | - |
| McGill Pain questionnaire – miscellaneous dimension | - | - | 8.54 | 4.17 | - | - | - |
| McGill Pain questionnaire – total score | - | - | 43.23 | 12.50 | - | - | - |
| Total widespread index (American College of Rheumatology criteria 2016)<sup>\$</sup> | .97 | 1.43 | 11.41 | 4.05 | **-12.705** | **31.770** | **<0.001** |
| Total symptom severity score (American College of Rheumatology criteria 2016)<sup>\$</sup> | 1.47 | 1.48 | 8.19 | 2.59 | **-11.860** | **40.412** | **<0.001** |
| TAS total score | 39.40 | 12.18 | 55.33 | 20.30 | **-3.545** | **41.663** | **<0.001** |
| HAMD total score | 1.33 | 1.99 | 14.67 | 6.52 | **-10.204** | **30.342** | **<0.001** |
| HAMA total score | 2.33 | 2.50 | 20.63 | 7.02 | **-12.837** | **31.887** | **<0.001** |

SD: standard deviation; df: degrees of freedom; VAS: visual analogue scale; FIQ: fibromyalgia impact questionnaire; TAS: Toronto alexithymia scale; HAMD: Hamilton depression rating scale; HAMA: Hamilton anxiety rating scale
<sup>\$</sup> Five healthy controls reported some pain during the interview, due to high physical activity in the previous days. None of the participant from the control group experienced any chronic pain condition

### Behavioural results

Behavioural results are presented in Supplementary Material (Table S1 and Figure S1). The fibromyalgia group reported significantly lower levels of emotional state during conditions involving negative emotions (attend, reappraise, suppress) compared to controls, and reported higher levels of arousal while attending negative emotions. The fibromyalgia group reported significantly higher levels of pain during all conditions compared to controls.

### Brain activation

The linear regression model indicated a significant association between brain activation during suppression of negative emotions and group (*pFWEc* = 0.042), but not with TAS total score or the group-by-TAS interaction. During the suppression of negative emotions, females with fibromyalgia showed significantly weaker activation in the left middle frontal gyrus compared to controls (see Table 3A and Figure 1).

**Table 3.**
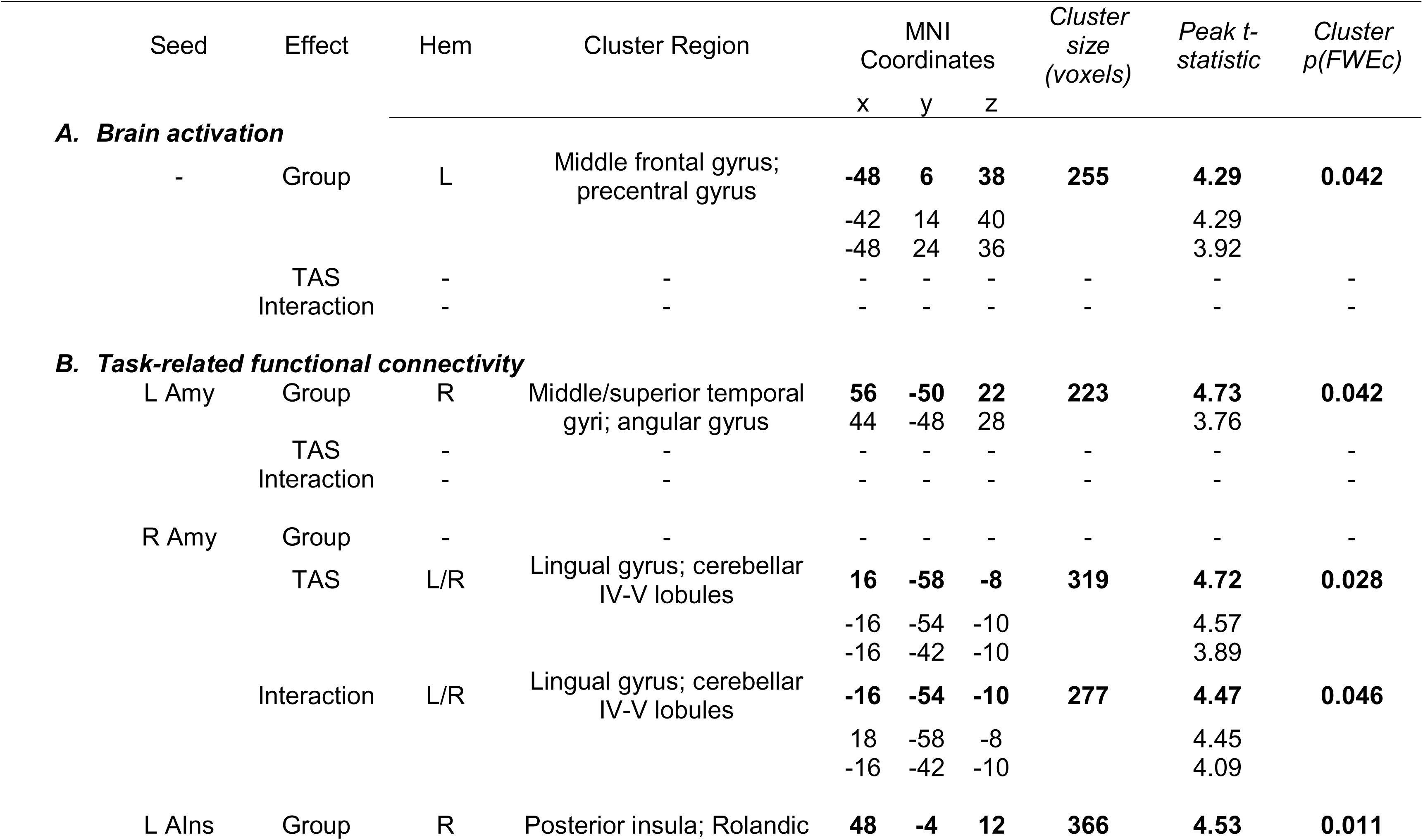
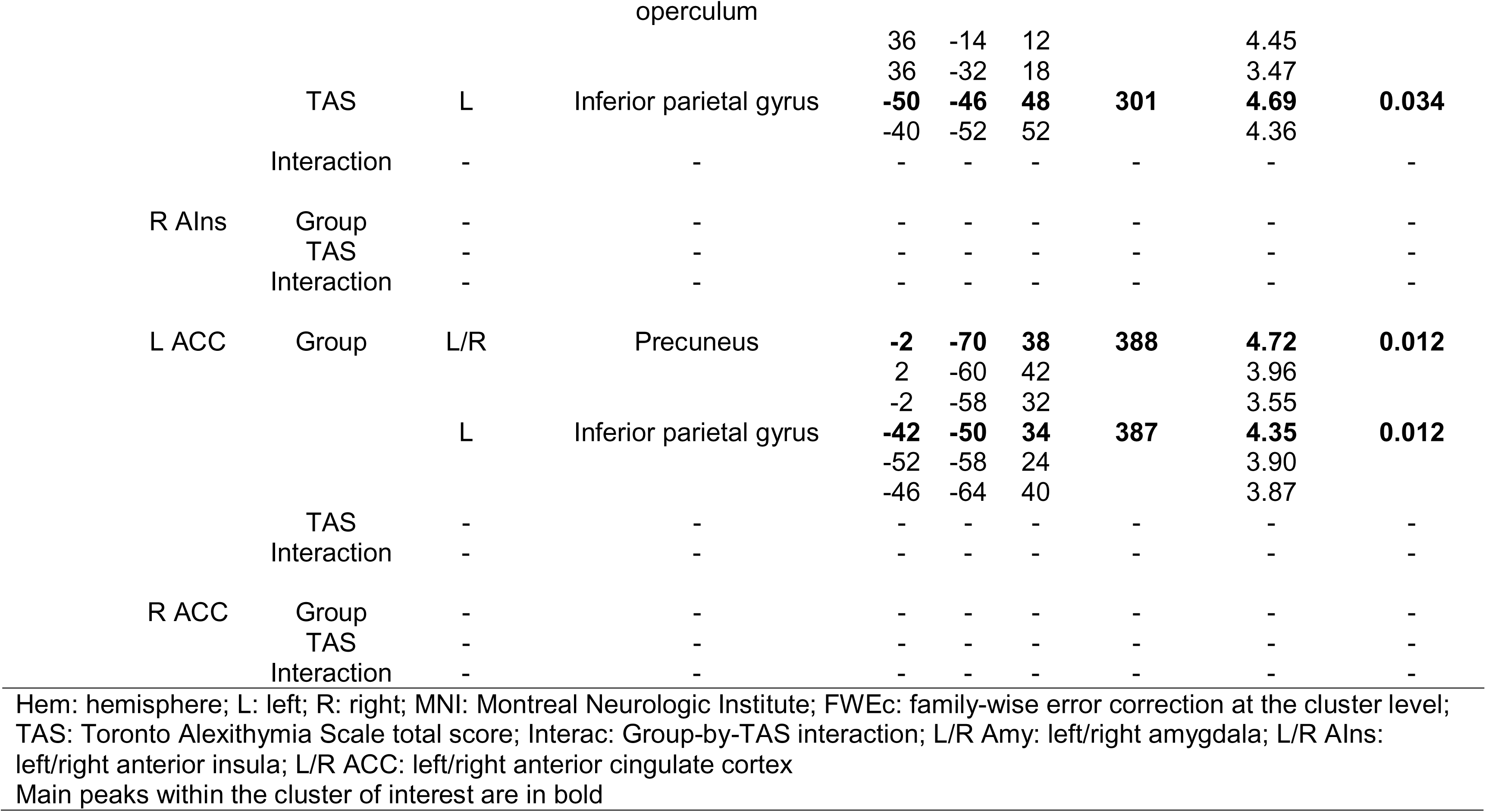
Peaks of clusters showing significant associations between Group, TAS total scores, or their interaction on (A) brain activation, and (B) task-related functional connectivity with the selected seed regions during suppression of negative emotions

**Figure 1.**
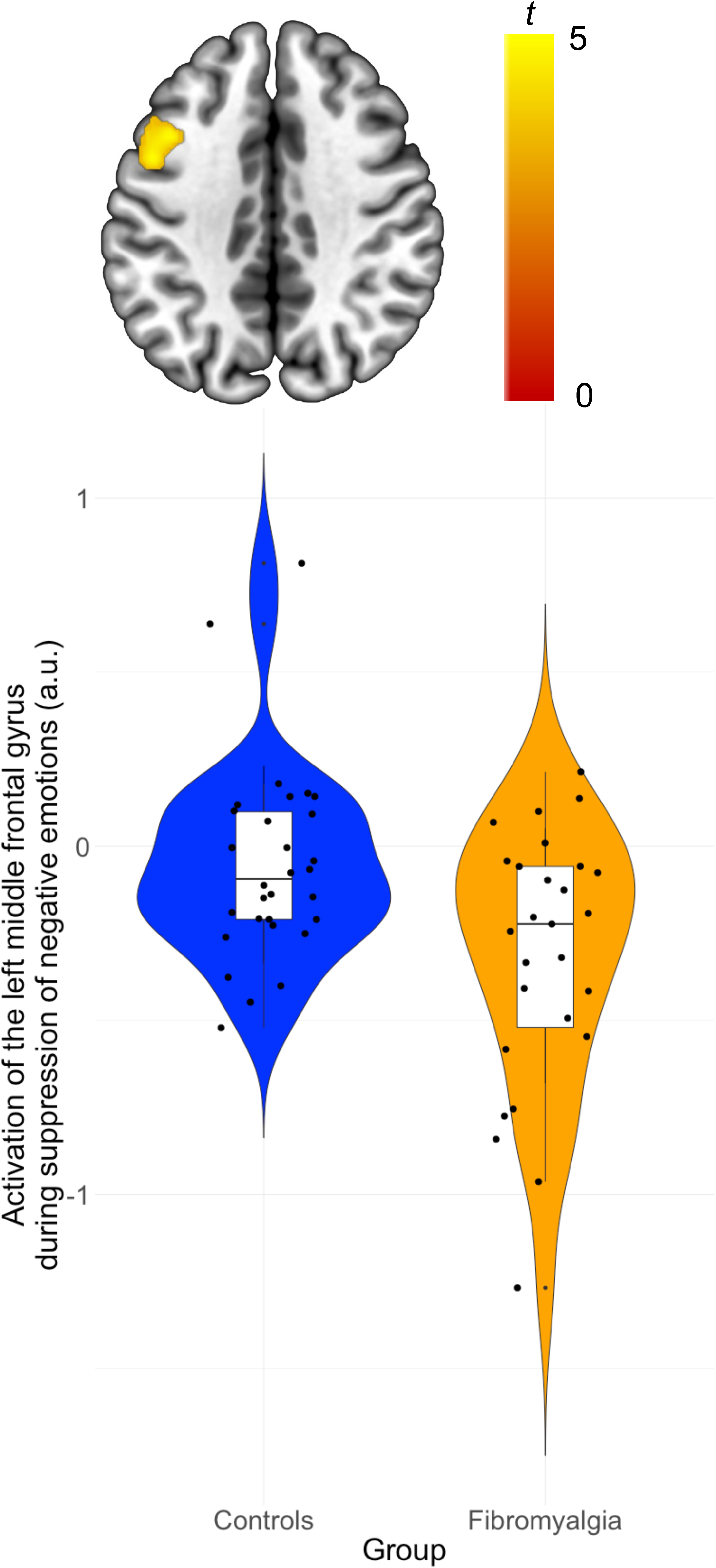
Group difference in brain activation during suppression of negative emotions. The group of females with fibromyalgia (in yellow) showed reduced activation in the left middle frontal gyrus compared to the group of pain free controls (in blue). Statistical significance was set at a voxel-wise threshold of *p*=0.001 uncorrected, to which a family-wise error (FWE) correction was applied on cluster statistics [*p(FWEc)* < 0.05]. Left is the left of the figure; a.u.: arbitrary units; colour-bar represents t-statistics

### Task-related functional connectivity

#### Left amygdala seed

The whole-brain multiple linear regression model showed a significant association between group and connectivity between the left amygdala seed and the right TPJ (*pFWEc* = 0.042) during suppression of negative emotions (see Table 3B and Figure 2A). Females with fibromyalgia showed weaker left amygdala-right TPJ connectivity compared to controls. Connectivity with the left amygdala seed was not significantly associated with TAS total scores or the group-by-TAS interaction.

**Figure 2.**
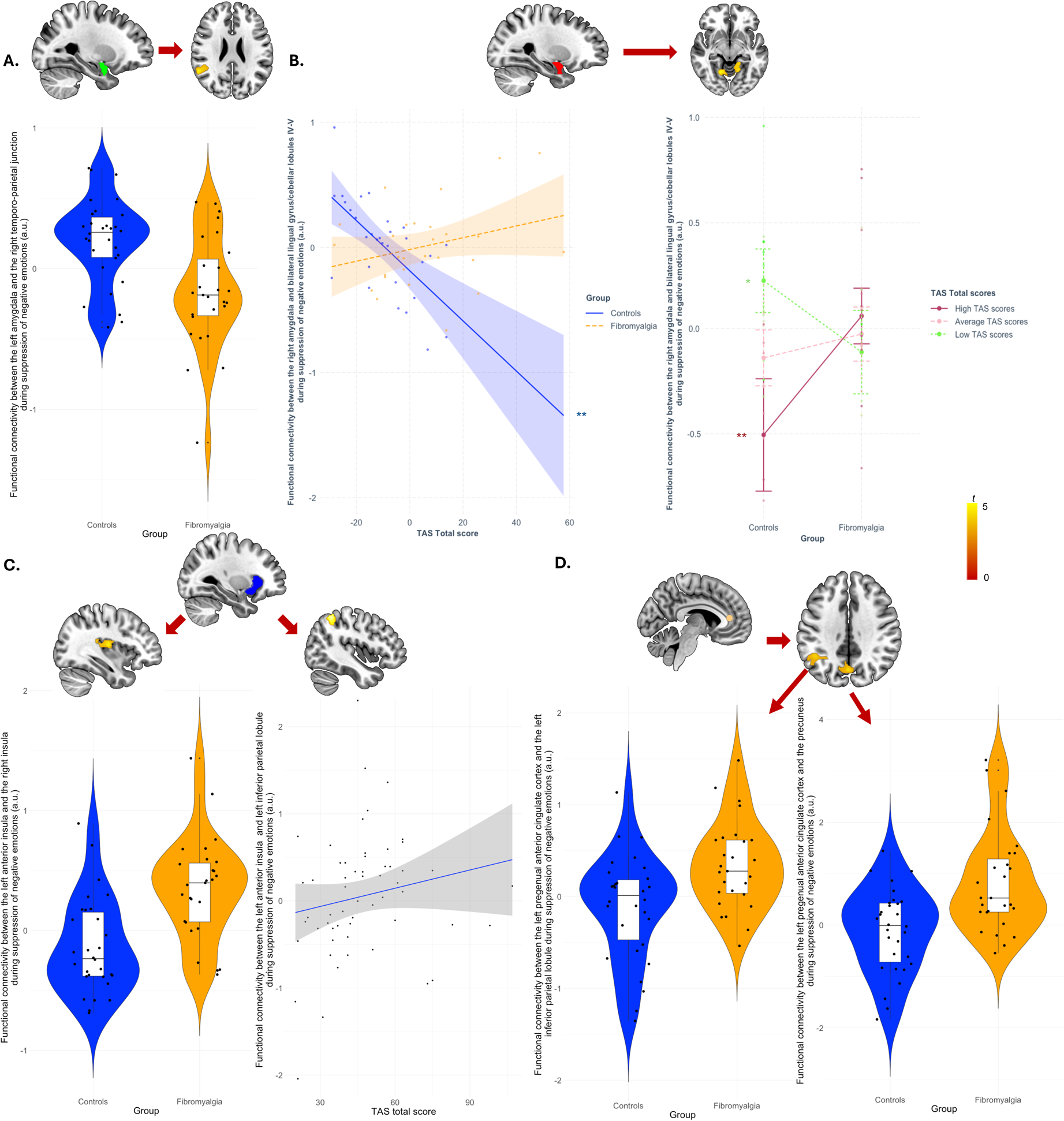
Group differences in task-related functional connectivity during suppression of negative emotions. (A) Compared to pain-free controls (in blue), the fibromyalgia group (in yellow) showed weaker connectivity between the left amygdala seed (in green) and the right temporo-temporal junction. (B) The relationship between Toronto alexithymia scale (TAS) total score and connectivity between the right amygdala seed (in red) and bilateral lingual gyri was moderated by group, with TAS total scores being significantly negatively associated with right amygdala-bilateral lingual gyri connectivity in controls (in blue) , but not in fibromyalgia (in yellow). In addition, the right amygdala-bilateral lingual gyri connectivity was significantly weaker in fibromyalgia compared to controls at lower TAS levels (in light green), but stronger at higher TAS levels (in maroon). Compared to controls (in yellow), the fibromyalgia group (in yellow) showed stronger connectivity between the (C) left anterior insula seed and the right posterior insula and (D) between the left anterior cingulate cortex (ACC) seed (in light pink) and the bilateral precuneus and the left inferior parietal lobule. In addition, connectivity between the left anterior insula and the left inferior parietal lobule was significantly and positively associated with TAS total scores. Statistical significance was set at a voxel-wise threshold of *p*=0.001 uncorrected, to which a family-wise error (FWE) correction was applied on cluster statistics [*p(FWEc)* < 0.05]. Left is the left of the figure; a.u.: arbitrary units; colour-bar represents *t*-statistics; \**p* < 0.05; \*\**p* < 0.01

#### Right amygdala seed

The whole-brain multiple linear regression model showed a significant association between the group-by-TAS interaction (*pFWEc* = 0.046) and TAS total scores (*pFWEc* = 0.028), and connectivity between the right amygdala seed and the bilateral lingual gyri during suppression of negative emotions (see Table 3B and Figure 2B). The first moderation analysis (group as moderator) showed that TAS total scores were significantly and negatively associated with right amygdala-bilateral lingual gyri connectivity during suppression on negative emotions in controls (*b* = -0.020, *se* = 0.006, *t* = -3.145, *p* = 0.003) but not in females with fibromyalgia (*b* = 0.005, *se* = 0.004, *t* = 1.089, *p* = 0.281). The second moderation analysis (TAS total score as moderator) showed that right amygdala-bilateral lingual gyri connectivity was significantly different between the groups at low (*b* = -0.338, *se* = 0.138, *t* = -2.444, *p* = 0.018; controls > fibromyalgia) and high (*b* = 0.564, *se* = 0.197, *t* = 2.865, *p* = 0.006; fibromyalgia > controls), but not average levels of TAS total scores (*b* = 0.113, *se* = 0.096, *t* = 1.171, *p* = 0.247). There was no significant association between connectivity of the right amygdala seed and group.

#### Left anterior insula seed

The whole-brain multiple linear regression model showed a significant association between group connectivity between the left anterior insula seed and the right posterior insula (*pFWEc* = 0.011) during suppression of negative emotions (see Table 3B and Figure 2C). During suppression of negative emotions, females with fibromyalgia showed heightened left anterior insula-right posterior insula connectivity compared to controls. In addition, the TAS total scores were significantly and positively associated with connectivity between the left anterior insula and the left inferior parietal lobule across all participants, independently of group (*pFWEc* = 0.034). There was no significant association between connectivity of the left anterior insula seed and the group-by-TAS interaction.

#### Right anterior insula seed

There was no significant association between group, TAS total scores or their interaction and functional connectivity with the right anterior insula seed (see Table 3B).

#### Left ACC seed

The whole-brain multiple linear regression model showed a significant association between group and connectivity between the left ACC seed and bilateral precuneus (*pFWEc* = 0.012) and the left inferior parietal lobule (*pFWEc* = 0.012) during suppression of negative emotions (see Table 3B and Figure 2D). Females with fibromyalgia showed heightened left ACC-bilateral precuneus and left ACC-left inferior parietal lobule connectivity compared to controls. Connectivity of the left ACC seed was not significantly associated with TAS total scores and the group-by-TAS interaction.

#### Right ACC seed

There was no significant association between group, TAS total scores or their interaction and functional connectivity with the right ACC seed (see Table 3B).

## Discussion

This study examined the neural correlates of negative emotion suppression in women with fibromyalgia and the moderating role of alexithymia on brain activation and functional connectivity. Compared to controls, the fibromyalgia group showed weaker activation in the left middle frontal gyrus, reduced left amygdala–right TPJ connectivity, and heightened connectivity between the left anterior insula and right posterior insula, and between the left ACC and both the bilateral precuneus and left inferior parietal lobule. Group-by-alexithymia interactions were associated with right amygdala–bilateral lingual gyri connectivity, with opposite patterns in controls and fibromyalgia, and alexithymia across participants was positively associated with left anterior insula–left inferior parietal lobule connectivity.

Compared to controls, fibromyalgia participants exhibited reduced activation in the left middle frontal gyrus, a core part of the dlPFC, important for cognitive control and emotion regulation (Chen et al., 2023). Reduced dlPFC recruitment during suppression may reflect impaired top-down modulation of limbic responses, contributing to affective reactivity and limited behavioural flexibility in emotionally salient contexts (Ochsner et al., 2012). Strengthening dlPFC engagement through emotion-regulation focused psychological approaches (Norman-Nott et al., 2025; Norman-Nott et al., 2024) or neuromodulation such as repetitive transcranial magnetic stimulation (rTMS) (Zhou et al., 2024), may normalise dlPFC-related function and enhance adaptive emotional and behavioural responses in fibromyalgia.

Beyond regional activation, group differences emerged in task-related functional connectivity. Women with fibromyalgia exhibited reduced connectivity between the left amygdala and the right TPJ. While no fibromyalgia study has directly examined amygdala–TPJ coupling, reduced connectivity here may reflect a breakdown in integration of affective and higher-order social-cognitive processes. The TPJ supports functions such as self–other differentiation and mentalising (Abu-Akel & Shamay-Tsoory, 2011), and interacts with the amygdala to enable flexible interpretation and modulation of emotional experiences (Nejati et al., 2023; Pessoa, 2010). Impaired connectivity may hinder reappraisal or contextualisation of negative stimuli, which may contribute to the difficulties in emotion regulation in fibromyalgia (Cavicchioli et al., 2025). Participants with fibromyalgia also showed increased connectivity between the left anterior insula and right posterior insula, and between the left ACC and the precuneus and left inferior parietal lobule while suppressing negative emotions. These patterns suggest heightened interoceptive and self-referential monitoring, potentially underlying the hypervigilance toward bodily sensations reported in fibromyalgia (Todd et al., 2024). The posterior insula encodes bodily signals, while the anterior insula integrates these into conscious awareness (Craig, 2009). Hyperconnectivity between the anterior and posterior insula may amplify somatic sensations and their emotional salience (Fermin et al., 2023; Namkung et al., 2017). Similarly, increased ACC-precuneus coupling, core node of the default mode network, may reflect excessive self-referential processing and reduced disengagement from internal states during suppression (Johansson et al., 2024). Together, these findings align with models such as FITSS that emphasise maladaptive integration of interoceptive, salience, and self-focused networks (Pinto et al., 2023). They highlight specific network interactions, particularly between the salience and default-mode networks, that may underlie hypervigilance and regulatory inefficiency in fibromyalgia. Such patterns offer potential neural targets and candidate biomarkers for testing interventions to normalise connectivity and improve emotional and physical symptom regulation.

Alexithymic traits exerted a differential impact on functional connectivity among the groups. In controls, higher alexithymia scores correlated with weaker right amygdala–bilateral lingual gyri connectivity, regions implicated in visual imagery and emotional encoding (Pessoa, 2010), potentially reflecting reduced visual–emotional integration consistent with prior non-clinical findings (Reker et al., 2010). This association was absent in individuals with fibromyalgia, suggesting that, during suppression of negative emotions, alexithymia disrupts visual imagery components typically supporting emotion suppression. These alterations align with evidence linking alexithymia in somatic and affective disorders to heightened ACC recruitment, and weaker activation in interoceptive and self-referential regions such as the insula and precuneus, potentially reflecting diminished emotional insight and granularity (van der Velde et al., 2013). Across all participants, alexithymia was positively associated with stronger left anterior insula–left inferior parietal lobule connectivity, suggesting heightened self-focused processing consistent with maladaptive cognitive styles and prior evidence of altered connectivity between regulation regions and the inferior parietal lobule (Montolio et al., 2025). These patterns may help identify neural targets and behavioural markers for interventions to improve emotional awareness and regulation in alexithymia, particularly in fibromyalgia.

Together, the results highlight potential targets for intervention by revealing disrupted integration across emotional, interoceptive, and self-referential networks in fibromyalgia. Interventions enhancing interoceptive accuracy include mindfulness-based approaches (Arey et al., 2024; Heagney & Adams, 2024; Voss et al., 2023).

Emotional clarity can be improved through emotion labelling or affective education, effective in higher emotional awareness. Cognitive regulation can be strengthened via reappraisal training or acceptance-based strategies (Eastwood & Godfrey, 2024; Heagney & Adams, 2024). Emotion regulation–focused interventions also show benefits in chronic pain, with improvements in pain and affective outcomes (Norman-Nott et al., 2025; Norman-Nott et al., 2024), and neuromodulation, such as high frequency rTMS over the left dlPFC, showed analgesic effects across chronic pain trials and may normalise regulatory network function (Zhou et al., 2024). Home-based anodal tDCS over the left dorsolateral prefrontal cortex (left-DLPFC) can reduce aberrant ACC–insula connectivity, indicating a potential normalisation of maladaptive dynamics (Lopes Alves et al., 2024). By reducing hypervigilance to bodily signals and fostering engagement with emotional cues, these interventions, separately or in conjunction, may alleviate distress and improve emotion regulation in fibromyalgia, particularly with high alexithymia.

Several limitations should be noted. First, the study focused exclusively on suppression, limiting conclusions about broader emotion regulation difficulties in fibromyalgia. Other strategies, such as reappraisal, avoidance, or acceptance, may engage distinct neural systems (Monachesi et al., 2023; Schmitz et al., 2021), consistent with the maladaptive (avoidance, suppression) or adaptive (appraisal) nature of these strategies (Koechlin et al., 2018). Second, reliance on TAS total score incompletely captures alexithymia’s multidimensional structure. Newer tools like the Perth Alexithymia Questionnaire may offer more precise assessment in chronic pain (Aaron et al., 2025; Preece et al., 2024). Third, focusing solely on negative suppression overlooks regulation of positive affect and ambivalent states, also relevant in pain (Borg et al., 2018; Monachesi et al., 2023). Fourth, the sample comprised only women, restricts generalisability. Fifth, the high prevalence of post-traumatic stress disorder in fibromyalgia (Vidal et al., 2025), and overlapping salience, default mode, and limbic circuits in both conditions (Akiki et al., 2017; Zandvakili et al., 2020), may confound findings and warrant consideration in future research. Finally, the relatively small sample size limits the statistical power of this study, with connectivity results not surviving Bonferroni correction. Brain-wide association work shows reproducible brain–behaviour effects typically require much larger samples or optimised study designs (e.g., thousands) (Marek & Dosenbach, 2018). Recruiting such clinical populations remains challenging; only large consortia such as the Enhancing NeuroImaging Genetics through Meta-Analysis (ENIGMA)-Chronic Pain working group (Quidé et al., 2024) can achieve this. Finally, the cross-sectional design of this study precludes causal inference.

Taken together, the findings indicate that negative emotion suppression in fibromyalgia is characterised by disrupted coordination between emotional, interoceptive, and self-referential networks. Alexithymia further modulated some of these responses, amplifying interoceptive reactivity and limiting emotional clarity. Overall, the results support a model where altered salience and default mode network dynamics contribute to regulation difficulties in fibromyalgia, pointing to potential targets for intervention. Targeting these networks through neuromodulation, mindfulness, emotional awareness training, and cognitive regulation may reduce bodily hypervigilance and improve emotion regulation in fibromyalgia, particularly among individuals experiencing alexithymia.

## Acknowledgments

The authors would like to acknowledge the use of the ds004144 dataset, which is publicly accessible through Openneuro.org (doi:10.18112/openneuro.ds004144.v1.0.2) and described in Balducci et al., 2022 (doi: 10.1038/s41597-022-01677-9)

## Data availability

The data that support the findings are fully available from Openneuro.org (doi:10.18112/openneuro.ds004144.v1.0.2)

## Authors contributions

A.A.C.V contributed conceptualization, methodology, validation, writing of the original draft, review and editing. N. N-N contributed conceptualization, investigation, methodology, supervision, and review and editing. S.M.G. contributed conceptualization, data curation, formal analysis, investigation, methodology, project administration, resources, supervision, and review and editing. Y.Q. contributed conceptualization, data curation, formal analysis, investigation, methodology, validation, visualization, supervision, writing of the original draft, and review and editing.

