## Supplementary Material for "Alexithymia moderates brain function during suppression of negative emotions in fibromyalgia"

Amaia Aguirre de Cárcer Vidal^1,2,3^, Nell Norman-Nott^1,2^, Sylvia M. Gustin^1,2^, Yann Quidé^1,2,*^

^1^ NeuroRecovery Research Hub, The University of New South Wales (UNSW) Sydney, Sydney, NSW, Australia

^2^ Centre for Pain IMPACT, Neuroscience Research Australia, Randwick, NSW, Australia

^3^ School of Psychology and Counselling, Queensland University of Technology, Brisbane, QLD, Australia

*Corresponding author:

Dr Yann Quidé, NeuroRecovery Research Hub, School of Psychology, Biological Sciences (Biolink) building, Level 1, UNSW Sydney, NSW, 2052, Australia.

**Supplementary Methods**

*Images processing and denoising*

Image processing followed the CONN toolbox’s default pipeline (release 22.v2407; https://web.conn-toolb ox.org) (Whitfield-Gabrieli & Nieto-Castanon, 2012) for SPM12 (v7771; Wellcome Trust Centre for Neuroimaging, London, UK; http://www.fil.ion.ucl.ac.uk/spm) in Matlab r2024a (Mathworks Inc., Sherborn, MA, USA). Functional and anatomical data were pre-processed using a modular preprocessing pipeline including realignment with correction of susceptibility distortion interactions, slice timing correction, outlier detection, direct segmentation and MNI-space normalization, smoothing, and regression of temporal components. Functional data were realigned using SPM realign & unwarp procedure, where all scans were co-registered to a reference image (first scan of the first session) using a least squares approach and a 6 parameter (rigid body) transformation and resampled using b-spline interpolation to correct for motion and magnetic susceptibility interactions. Temporal misalignment between different slices of the functional data was corrected following SPM slice-timing correction (STC) procedure, using sinc temporal interpolation to resample each slice BOLD timeseries to a common mid-acquisition time. Potential outlier scans were identified using ART as acquisitions with framewise displacement above 2 mm or global BOLD signal changes above 9 standard deviations (Power et al., 2014), and a reference BOLD image was computed for each subject by averaging all scans excluding outliers. Functional and anatomical data were normalized into standard MNI space, segmented into grey matter, white matter, and CSF tissue classes, and resampled to 2 mm isotropic voxels following a direct normalization procedure using SPM unified segmentation and normalization algorithm with the default IXI-549 tissue probability map template. Functional data were smoothed using spatial convolution with a Gaussian kernel of 8 mm full width half maximum (FWHM). Last, components from were removed from the BOLD signal timeseries using a separate linear regression model at each individual voxel.

In addition, functional data were denoised using a standard denoising pipeline including the regression of potential confounding effects characterized by white matter timeseries (5 CompCor noise components), CSF timeseries (5 CompCor noise components), motion parameters and their first order derivatives (12 factors), and linear trends (2 factors) within each functional run, followed by high-pass frequency filtering of the BOLD timeseries above 0.008 Hz. CompCor (Behzadi et al., 2007) noise components within white matter and CSF were estimated by computing the average BOLD signal as well as the largest principal components orthogonal to the BOLD average, motion parameters within each subject's eroded segmentation masks.

**Supplementary Results**

***Behavioural results***

| Table S1. Behavioural results | | | | | | | | |
| --- | --- | --- | --- | --- | --- | --- | --- | --- |
|  | Controls (n=30) | | Fibromyalgia (N=27) | | Statistics | | | |
|  | Mean | SD | Mean | SD | Welch | df | *p*-value | |
| **VAS Arousal** |  |  |  |  |  |  |  | |
| Attend neutral | 5.424 | 0.983 | 4.938 | 1.361 | 1.648 | 56.343 | | 0.105 |
| Attend positive | 5.596 | 1.260 | 5.583 | 1.830 | 0.032 | 54.844 | | 0.974 |
| Attend negative | **5.586** | **1.225** | **6.458** | **1.777** | **-2.298** | **54.854** | | **0.025** |
| Reappraise positive | 5.657 | 1.324 | 5.969 | 1.658 | -0.837 | 59.253 | | 0.406 |
| Reappraise negative | 5.667 | 1.164 | 5.844 | 1.698 | -0.489 | 54.689 | | 0.627 |
| Suppress positive | 5.475 | 1.261 | 5.615 | 1.245 | -0.450 | 62.979 | | 0.654 |
| Suppress negative | 5.667 | 1.310 | 5.979 | 1.788 | -0.802 | 56.765 | | 0.426 |
| **VAS Emotional State** |  |  |  |  |  |  |  | |
| Attend neutral | 5.697 | 1.245 | 5.313 | 1.230 | 1.252 | 62.978 | 0.215 | |
| Attend positive | 7.071 | 1.592 | 6.375 | 1.656 | 1.726 | 62.683 | 0.089 | |
| Attend negative | **3.818** | **1.852** | **2.750** | **1.737** | **2.399** | **62.932** | **0.019** | |
| Reappraise positive | 7.657 | 1.464 | 7.219 | 1.556 | 1.168 | 62.470 | 0.247 | |
| Reappraise negative | **4.535** | **1.648** | **3.490** | **1.604** | **2.593** | **62.999** | **0.012** | |
| Suppress positive | 6.323 | 1.494 | 5.906 | 1.331 | 1.189 | 62.560 | 0.239 | |
| Suppress negative | **4.273** | **1.519** | **3.135** | **1.800** | **2.749** | **60.611** | **0.008** | |
| **VAS Pain** |  |  |  |  |  |  |  | |
| Attend neutral | **0.970** | **1.690** | **5.240** | **2.438** | **-8.182** | **55.045** | **<0.001** | |
| Attend positive | **0.778** | **1.363** | **5.073** | **2.628** | **-8.232** | **46.237** | **<0.001** | |
| Attend negative | **0.859** | **1.850** | **6.313** | **2.444** | **-10.122** | **57.759** | **<0.001** | |
| Reappraise positive | **0.838** | **1.720** | **5.021** | **2.685** | **-7.452** | **52.523** | **<0.001** | |
| Reappraise negative | **0.960** | **2.061** | **5.896** | **2.410** | **-8.862** | **60.891** | **<0.001** | |
| Suppress positive | **0.596** | **1.416** | **5.385** | **2.471** | **-9.547** | **49.062** | **<0.001** | |
| Suppress negative | **0.980** | **1.639** | **5.813** | **2.445** | **-9.331** | **53.979** | **<0.001** | |
| VAS: visual analogue scale score; SD: standard deviation; df: degrees of freedom | | | | | | | | |

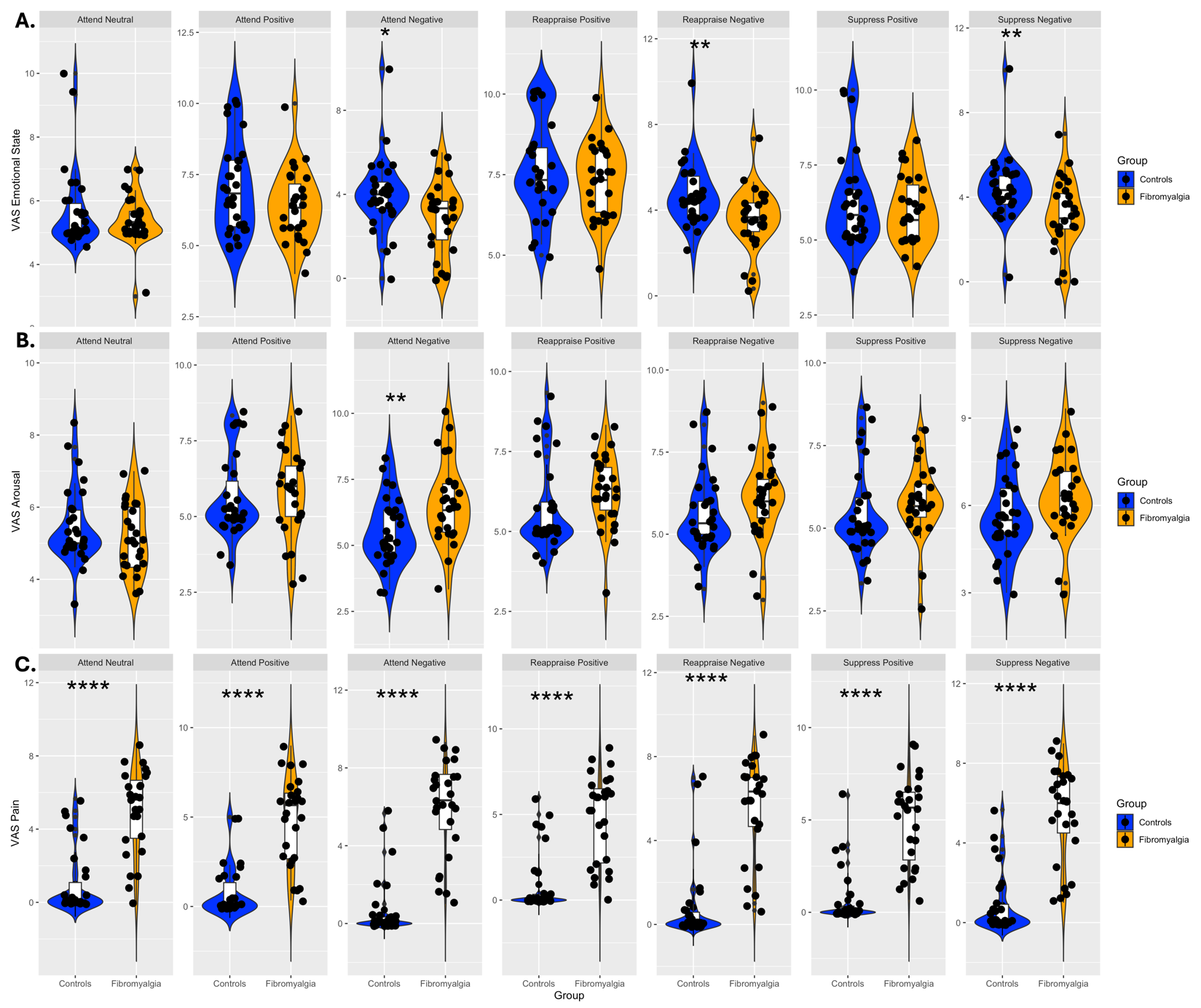

**Figure S1. Representation of the behavioural results**

Group differences between the fibromyalgia (orange on the right side) and control groups (in blue, on the left) on the visual analogue scales (VAS) for (A) arousal, (B) emotional state, and (C) pain intensity for all task conditions, from left to right: attend neutral, attend positive, attend negative, reappraise positive, reappraise negative, suppress positive and suppress negative emotions.

**p*< 0.05, ***p* < 0.01, *****p*<0.0001
